# Safety and tolerance of infant formulas containing probiotics: a multicenter, randomized, controlled trial in healthy term infants

**DOI:** 10.1101/2025.04.29.25326625

**Authors:** M. Mitra, A. Petit-Jean, R. Agrawal, U. Vaidya, S. Ganguly, N. P. Hays

## Abstract

Several studies have documented the safety and tolerance of probiotics in infants, however, most studies are conducted with supplement. This randomized, double-blind, multicenter trial evaluated growth of healthy term infants fed infant formula supplemented with *Limosilactobacillus reuteri DSM17938* (*L. reuteri*; n=92) or *Bifidobacterium lactis* CNCM I-3446 *(B. lactis*; n=92) or the same formula without probiotics (n=95). Mixed feeding with breast milk was allowed in each group. Exclusively breastfed infants (n=100) were included for reference. Non-inferiority in weight gain (margin −3 g/day) from enrollment to age 6 months was the primary outcome. Length, BMI, head circumference, and WHO *z*-scores from birth to 12 months were assessed, as was digestive tolerance, and, in a subset of infants, urinary D-lactate parameters and stool microbiota composition. Of 279 infants randomized, 256 completed the study. The mean difference in weight gain between each probiotic group and the standard group at age 6 months was −0.378 g/day (97.5% confidence interval [CI], −1.541, 0.776; *P*<0.001) for *L. reuteri* and −1.724 g/day (97.5% CI, −2.845, −0.603; *P*=0.005) for *B. lactis*, indicating non-inferior growth. Anthropometric *z*-scores were not significantly different between any of the formulas, over the entire study, except for a slightly lower weight-for-age *z*-score in the *B. lactis* vs. standard group at 8 months (*P*=0.034). No differences in adverse events or urinary D-lactate levels were observed. Significantly higher fecal Lactobacilli counts were observed with *L. reuteri* supplementation (7.0 log10[CFU/g]; n=24) compared with standard formula (6.2 log10[CFU/g]; n=21). Parent-reported digestive tolerance symptoms were similar among the formula groups and comparable to the breastfed group. Weight gain from enrollment to age 6 months in Indian infants fed formula containing probiotics *L. reuteri DSM17938* or *B. lactis* CNCM I-3446 was non-inferior versus infants fed the same formula without probiotics. Both probiotic formulas were safe and well-tolerated.

**Trial registration:** CTRI/2010/091/001111 (Clinical Trials Registry - India; www.ctri.nic.in)

## 1. Introduction

A greater understanding of human milk composition and the differences between breastfed and formula-fed infants have provided opportunities to continuously improve formulas for infants unable to be breastfeed. One major difference between breastfed and formula-fed infants is a difference in gut microbiota composition, which may result in a lower incidence of infections and morbidity in breastfed infants (Odiase *et al*., 2023; Stuebe, 2009).

In breastfed full-term infants, *lactobacilli* and *bifidobacterium* begin to appear in the stool by the third day of life (Yao *et al*., 2021). *Bifidobacterium* then becomes the dominant organism, representing 80% to 90% of the gut ecosystem. Formula-fed infants are colonized by a more diverse microbiota (Yang *et al*., 2016), These differences in the composition of the intestinal microbiota appear to be related to the more acidic environment of the gut in breastfed children (Duar *et al*., 2020), which tends to favor the predominance of bifidobacteria and inhibit the growth of potentially pathogenic bacteria (Gibson and Roberfroid, 1995).

Adding probiotics to infant formulas is an important innovation designed to improve the intestinal microbiota of formula-fed infants. The addition of specific probiotics (e.g., strains of Bifidobacteria) may also help improve the growth of infants, particularly in vulnerable populations (Steenhout *et al*., 2009). Several probiotics have been evaluated in randomized, controlled, double-blind clinical trials of infants and toddlers and have been shown to be safe and well-tolerated (Chouraqui *et al*., 2008; Gibson *et al*., 2009; Radke *et al*., 2017; Shen *et al*., 2024; Steenhout *et al*., 2009; Weizman *et al*., 2005). Probiotics such as *Bifidobacterium animalis* subspecies *lactis* CNCM I-3446 (*B. lactis*) have been shown to reduce the incidence of diarrhea (Chouraqui *et al*., 2004; Saavedra *et al*., 1994), diaper rash (Hertiš Petek *et al*., 2022; Saavedra *et al*., 1998), and hard stools (Saavedra *et al*., 1998) in a number of infant and toddler trials, while oral administration of *Limosilactobacillus reuteri* DSM 17938 (*L. reuteri*), has been reported to reduce colic (Savino *et al*., 2007), improve gastric emptying (Indrio *et al*., 2008; Indrio *et al*., 2011), and reduce regurgitation (Indrio *et al*., 2008; Indrio *et al*., 2011) in infants. Two other probiotics commonly added to infant formulas include *Bifidobacterium longum* ATCC BAA-999 and *Lacticaseibacillus rhamnosus* CGMCC 1.3724. Both have been shown to be safe and well tolerated as supplements in infant formulas (Berseth *et al*., 2024; Hascoet *et al*., 2011; Shulman *et al*., 2022). Evidence suggests that *Lacticaseibacillus rhamnosus* GG may help prevent atopic dermatitis and improve immune responses (Kaila *et al*., 1992; Kalliomaki *et al*., 2001). Because bifidobacteria act in the colon and lactobacilli act in the small intestine (Bennet *et al*., 1992), a mixture of both probiotics could provide benefit throughout the gut. Such a combination has previously been shown to be safe and well tolerated (Chouraqui *et al*., 2008).

Although the safety of infant formulas containing probiotics has been demonstrated in a variety of infant populations (Gibson *et al*., 2009; Papagaroufalis *et al*., 2014; Soh *et al*., 2009; Underwood *et al*., 2013; Weizman *et al*., 2005), safety and tolerance data in Indian infants are lacking. Evaluating the safety of infant formula with regard to growth is particularly important in India, a region in which growth failure remains a persistent challenge (Lundeen *et al*., 2014; Saji *et al*., 2024). Consequently, the primary objective of the present study was to evaluate weight gain in infants in India who received one of two infant formulas supplemented with probiotics, either *L. reuteri* or *B. lactis*, or the same infant formula without probiotics. Secondary objectives included the assessment of the impact of these probiotic-supplemented formulas on GI tolerance, adverse events (AEs), as well as fecal microbiota and urinary D-lactate.

## 2. Materials and Methods

### Study design

This was a prospective, multicenter, randomized, double-blind study to evaluate the safety and tolerance of infant formulas containing probiotics *Limosilactobacillus reuteri* (*L. reuteri DSM17938*) or *Bifidobacterium lactis (B. lactis* CNCM I-3446) in healthy term infants in India.

The study was conducted between August 2010 and July 2012 at 5 centers in India (CTRI/2010/091/001111 [Clinical Trials Registry - India; www.ctri.nic.in]). The study protocol and informed consent form were reviewed and approved by an Independent Ethics Committee at each center. Written informed consent was obtained from the infants’ parents or legal guardian prior to enrollment.

### Population

The study population consisted of healthy, full-term infants ≥2 weeks to 3 months of age at enrollment who required mixed feeding (i.e., breast milk and formula in any proportion) due to partial lactation failure or informed choice by the mother. Eligible infants had a gestational age of 37-42 weeks and a birth weight of 2500-4500g. Exclusion criteria included infants from an HIV-positive mother or mother with gestational diabetes; any significant illness or malformation that could affect normal growth; and re-hospitalization prior to enrollment for more than 2 days (except those re-hospitalized due to jaundice). Infants were openly allocated to either exclusive breastfeeding or mixed feeding groups. Within the mixed-fed group, blinded random allocation, stratified by sex and center, was conducted to assign infants to one of the three study formulas (randomization lists were produced using R version 2.6.1).

### Study formulas

Mixed-fed infants were randomized in a 1:1:1 ratio to one of three feeding groups: mixed feeding with infant infant formula containing the probiotic *L. reuteri* DSM17938 (2 × 10^7^ cfu/g of powder, equivalent to a daily intake of 2.4 × 10^9^ cfu; the *L. reuteri* group); mixed feeding with infant formula containing the probiotic *B. lactis* CNCM I-3446 (1.8 × 10^7^ cfu/g of powder, equivalent to a daily intake of 2.2 × 10^9^ cfu; the *B. lactis* group); or mixed feeding with a standard control formula not containing probiotics (the standard group). Each infant formula contained adequate amounts of energy (670 kcal/L), protein (2.4 g/100 kcal, casein/whey ratio of 60:40), and other nutrients to support the normal growth of healthy infants. Parents/caregivers were instructed to follow guidelines printed on the label regarding appropriate volumes of formula to be offered per day. Breastfed infants received exclusive breast milk until the end of the 6^th^ month of age. For infants stopping breastfeeding during the 5^th^ and 6^th^ month of age, aa infant formula without probiotics was provided until the end of 6 months. No other food was given to infants during the first 6 months of life. However, complementary foods other than the study formulas were permitted to be introduced gradually after age 6 months.

When these enrolled mixed-fed infants reached 7 months of age, they received the following assigned follow-up formulas until age 12 months: the *L. reuteri* group received follow-up formula supplemented with *L. reuteri* (1.9 × 10^7^ cfu/g of powder, equivalent to a daily intake of 2.1 × 10^9^ cfu; the *B. lactis* group received follow-up formula supplemented with *L. rhamnosus* CGMCC 1.3724 (1.5 × 10^7^ cfu/g of powder, equivalent to a daily intake of 1.7 × 10^9^ cfu) and *B. longum* ATCC BAA-999 (7.8 × 10^6^ cfu/g of powder, equivalent to a daily intake of 8.8 × 10^8^ cfu) and the standard group received follow-up formula without probiotics. Breastfed infants continued to receive exclusive breast milk or mixed feeding with follow-up formula with probiotics *L. rhamnosus* and *B. longum*, according to the parent’s choice.

The investigators, study staff, and parents were blinded to the identities of the formulas.

### Observation period

Infant demographic and anthropometric data were collected from birth records and at the enrollment visit. Enrollment was conducted between 2 weeks and 3 months after birth. Clinic visits were scheduled at 1.5, 2, 2.5, 3.5, 4.5, 6, 8, 10, and 12 months (visits 1-9) to dispense formula, collect anthropometry data, and record AEs (since the enrollment age could vary, the first three visits did not apply to some subjects). Parents were asked to record the feeding history of their infants for 3 consecutive days prior to each visit, up until 6 months. Feeding history included volume of formula (or breast milk) consumed and the number of formula feedings and breastfeeds per day. Parents/caregivers also recorded gastrointestinal (GI) tolerance for 3 consecutive days immediately prior to each of the clinic visits until 6 months.

### Outcomes

#### Growth and other anthropometric measurements

The primary outcome for this study was weight gain (g/day) from baseline (visit 0) to 6 months of age (visit 6). The primary hypothesis of the study was that the mean weight gain of infants fed a infant formula containing either *L. reuteri* or *B. lactis* up to 6 months of age is not inferior to the growth observed in infants fed with a infant formula without probiotics.

Infant weight was assessed at each clinic visit and rounded to the nearest 10 g. Infants were weighed without clothing or nappy/diaper on calibrated electronic weighing scales, with the same scales used for all infants at all visits.

Other anthropometric parameters assessed from birth to 12 months included length, body mass index (BMI), head circumference gains, and corresponding WHO *z*-scores. Recumbent length was measured at each visit in centimeters (±1 mm) using a standardized length board. Head circumference (±1 mm) was measured approximately 2.5 centimeters above the eyebrows, directly over the largest circumference of the skull.

#### Digestive tolerance

Digestive tolerance was determined by the investigator at each visit up to 6 months of age based on parents’ reports of stool consistency and frequency (number of stools per 24 hours), spitting up (no/sometimes/often), vomiting (no/sometimes/often), flatulence (no/sometimes/often), irritability/fussiness (yes/no, and if so for how many hours), and sleeping habits (number of times waking during the night, longest period of uninterrupted sleep). All information on the child’s stooling pattern and other GI tolerance symptoms was recorded in diaries by parents/caregivers for the 3 consecutive days immediately prior to each of the clinic visits until age 6 months. The investigator confirmed the child’s digestive tolerance based on a review of these diaries at each study visit.

#### Adverse events

Adverse events were collected at each visit throughout the entire 12-month study period, with a focus on diarrhea and upper respiratory tract infections (including bronchiolitis and otitis media). Any episodes of illness occurring since the last visit were recorded as AEs. Conditions associated with morbidity were predefined by investigators to help caregivers assess these events, including diarrhea (defined as 3 or more loose or watery stools per day, with an episode of diarrhea considered to have ended once there have been 2 consecutive non-watery stools, or no stools for 24 hours); respiratory symptoms (defined as runny nose or chronic cough recorded on a scale from 0 to 3 [0:absent; 1:mild; 2:moderate; 3:severe); fever (defined as when the infant’s temperature was above 38°C, reaching 38.5°C at least once during the last 24 hours); eczema (recorded on a scale from 0 to 3, similar to the one described for respiratory symptoms) and constipation (defined as when the infant does not pass a stool for ≥3 consecutive days, with stools that are hard, dry, and painful to pass).

#### Stool microbiota composition

At 4.5 months of age, a stool sample was obtained at home by the parents/caregivers in a subset of approximately 30 subjects per group in appropriate conditions, defined as less than 15 hours between stool emission and stool processing at the laboratory. *Lactobacilli, Bifidobacterium, Clostridium perfringens, Enterobacteriaceae, Bacteroides/Pevotella*, and total bacteria populations were analyzed by fluorescent *in situ* hybridization (BioVisible, Groningen, The Netherlands), and *B. lactis and L. reuteri* detection by plating and polymerase chain reaction methods (Advanced Analytical Technologies, Piacenza, Italy).

#### Urinary D-lactate

Following 2 months of formula- or breast-feeding, urinary samples were collected in a subset of approximately 15 subjects per group to analyze creatinine, D-lactate, L-lactate, % of D-lactate/L-lactate, and sum of D-lactate and L-lactate. Infants were included in this group if their parents or caregivers were willing and able to provide urine samples on their behalf. Samples were collected by study staff using a sterile urine collection bag approximately 2 hours after the last feed. Urine samples were aliquoted into cryotubes, frozen, and shipped on dry ice to a central laboratory for analysis (Clinical Chemistry Laboratory, Centre Hospitalier Universitaire Vaudois, Lausanne, Switzerland). Both D- and L-lactate concentrations were normalized per mole creatinine (mmol lactate/mol creatinine).

### Statistical methods

The sample size required to evaluate non-inferiority of the primary outcome (weight gain, g/day), based on a non-inferiority margin of −3.0 g/day and standard deviation (SD) for weight gain of 6.1 g/day (Barclay *et al*., 2003), was calculated as 79 infants per group. Since there were two primary comparisons (*L. reuteri* vs. standard formula and *B. lactis* vs. standard formula), Bonferroni’s adjustment was used to control the over-all type-1 error rate. With a type I error of 0.0125, a non-inferiority margin of −3 g/day, and SD of 6.1, a one-sided, two-sample z-test would require a sample size of 79 in each arm to detect non-inferiority with 80% power. Statistical analyses were performed using SAS Version 9.2 for Windows (Cary, North Carolina, US). The primary analysis was based on an intent-to-treat (ITT) approach. To compare the two probiotic formulas with the standard formula without probiotics with regard to weight gain from enrollment to 6 months of age (primary analysis), a mixed model with repeated visits was the choice of model because it provides a useful strategy to handle missing data due to dropout/loss-to-follow-up. The model was adjusted for sex, center, and birth weight. The study was considered a success and non-inferiority was accepted if the lower bound of the two-sided 97.5% confidence interval (CI) of the mean difference was greater than −3 g/day for each of the 2 probiotic formula groups.

Differences in other anthropometric parameters between the two probiotic formulas and the standard formula were analyzed in both the ITT population using a repeated measures model adjusted for sex, center, and parameter value at birth. Stool frequency counts were aggregated to one total value of counts (i.e., all stools were reported for each subject and divided by the number of available days). Two-sided superiority testing for treatment differences between groups was assessed by Poisson regression on the stool count per day. Percentage of stools characterized as hard, formed, soft and liquid stools, was defined as number of stools with certain consistency divided by the total number of stools. For each stool consistency category, logistic regression was used to assess between-group superiority. Other GI tolerance symptoms such as spitting up, vomiting and sleeping data were assessed by Poisson regression, while flatulence and irritability/fussiness data were assessed by logistic regression.

Gut microbiota composition was expressed in log base 10 and was analyzed by ANOVA. Parametric statistics were used and values below the detection limit were imputed to the detection limit value defined as 9.8 ^e5^ bacteria/g of feces. Urinary lactate data was adjusted by log transformation (log base e) and analyzed using the same model as for gut microbiota composition. Finally, the incidences of AEs were compared between groups by Chi-Square test. All gut microbiota data produced in the present study are available upon reasonable request to the authors.

## 3. Results

### Study population

Two hundred seventy-nine mixed-fed infants were randomized to the *L. reuteri*-supplemented infant formula (*L. reuteri*, n = 92), *B. lactis*-supplemented infant formula (*B. lactis*, n = 92), or the standard infant formula without any probiotics (standard, n = 95). One hundred breastfed infants were also enrolled. A total of 256 mixed-fed infants and 95 breastfed infants completed the trial (**Figure 1**).

**Figure 1.**
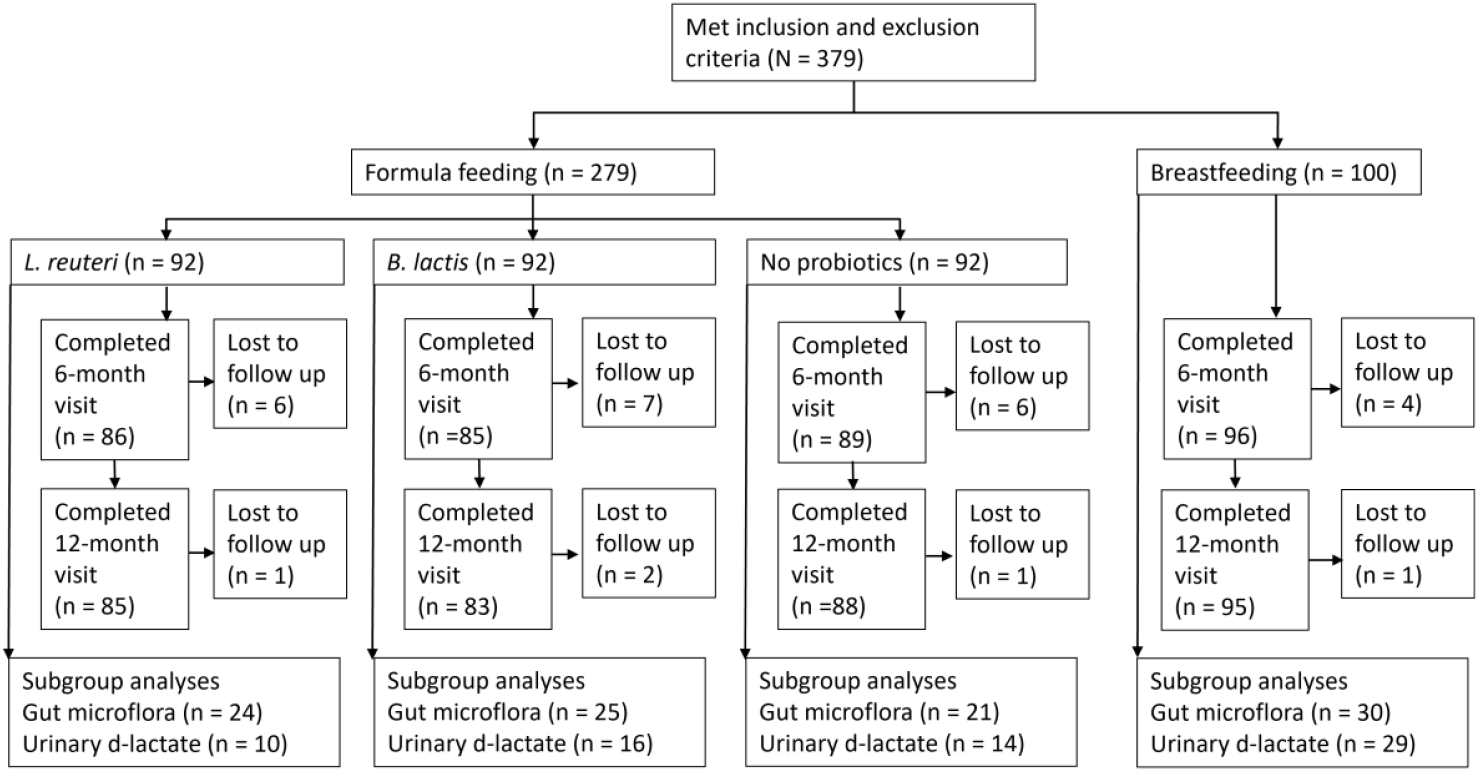
Subject disposition.

Of the 23 mixed-fed infants who did not complete the trial, all were lost to follow-up. No infants were reported to have discontinued the study due to AEs or tolerability concerns. Demographic and anthropometric characteristics at birth and at baseline were generally similar among groups (**Table 1**). Mean age at randomization ranged from 53 days among breastfed infants to 61 days among infants assigned to the *L. reuteri* group.

**Table 1.** Baseline demographics and anthropometric characteristics.

|  | Randomized Mixed-Fed Groups |  |  | Non-randomized Reference Group |
| --- | --- | --- | --- | --- |
|  | *L. reuteri*<sup>a</sup><br>(n = 92) | *B. lactis*<sup>b</sup><br>(n = 92) | Standard<sup>c</sup><br>(n = 95) | Breastfed<br>(n = 100) |
| **Demographics** |  |  |  |  |
| Age at baseline, days | 61.2 ± 20.7 | 55.7 ± 22.6 | 59.1 ± 20.9 | 52.7 ± 25.6 |
| Girls, % | 52.2 | 46.7 | 49.5 | 43.0 |
| **Anthropometric measurements at baseline<sup>d</sup>** |  |  |  |  |
| Weight, kg | 4.42 ± 0.94 | 4.30 ± 0.95 | 4.44 ± 0.95 | 4.53 ± 0.98 |
| Length, cm | 55.6 ± 3.5 | 54.7 ± 4.0 | 55.5 ± 3.8 | 56.0 ± 3.9 |
| Body mass index, kg/m<sup>2</sup> | 14.2 ± 2.3 | 14.2 ± 2.1 | 14.3 ± 2.1 | 14.3 ± 1.9 |
| Head circumference, cm | 37.2 ± 1.9 | 37.3 ± 1.9 | 37.3 ± 1.8 | 37.2 ± 2.0 |
| **Anthropometric measurements at birth** |  |  |  |  |
| Weight, kg | 2.93 ± 0.34 | 2.95 ± 0.33 | 2.93 ± 0.34 | 2.99 ± 0.35 |
| Length, cm | 47.4 ± 3.2 | 47.5 ± 2.1 | 47.6 ± 2.5 | 48.5 ± 3.5 |
| Head circumference, cm | 34.1 ± 2.9 | 33.8 ± 1.1 | 33.3 ± 1.1 | 34.3 ± 2.1 |

Mean ± SD unless otherwise noted; <sup>a</sup>Infant formula with *L. reuteri*; <sup>b</sup>Infant formula with *B. lactis*; <sup>c</sup>Infant formula without probiotics; <sup>d</sup>Pre-randomization: the infants' ages varied between 14 days and 3 months of age when the measurements were recorded.

### Weight gain and growth

The mean difference in weight gain between the two probiotic supplement groups and the standard group at 6 months of age was −0.378 g/day (97.5% CI, −1.541, 0.776) with *L. reuteri* and −1.724 g/day (97.5% CI, −2.845, −0.603) with *B. lactis*. Because the lower bound of the 97.5% CI in both comparisons was greater than −3 g/d, both test formulas demonstrated non-inferiority in weight gain velocity compared to the standard formula. The mean (SD) weight gain (g/day) was 22.73 (5.47), 20.93 (5.42) and 21.73 (5.42) with the *L. reuteri*-supplemented formula, *B. lactis*-supplemented formula, and the standard formula, respectively.

Body length, BMI, and head circumference were comparable among the 3 mixed-fed groups at 6 months and 12 months of age. Compared to the breastfed group, body length was slightly but significantly greater in each mixed-fed group (*P* ≤ 0.012 for all comparisons) at 12 months of age. No significant differences in BMI between breastfed and mixed-fed groups were observed at 6 or 12 months. Head circumference was slightly but significantly greater in the *B. lactis* group than in the breastfed group at 6 months of age (*P* = 0.035) and at 12 months of age (*P* = 0.002). Head circumference was also statistically greater in the standard formula group than in the breastfed group at 12 months of age (*P* = 0.004).

Changes in weight-for age, length-for age, and head circumference-for-age z-scores are shown in **Figure 2**. At 6 months of age, weight-for-age z-scores were slightly higher in the *L. reuteri* group compared to the *B. lactis* group (*P* = 0.026). No differences between mixed-fed groups were noted at 12 months of age. Compared to the breastfed group, weight-for-age z-scores were significantly higher at 6 months of age in the *L. reuteri* group (*P* < 0.001) and the standard formula groups (*P* = 0.002) and at 12 months of age in all 3 mixed-fed groups (*P* ≤ 0.022 for all comparisons). No significant differences in length-for-age or head circumference-for-age z-scores between mixed-fed groups were noted at 6 or 12 months of age. However, compared to the breastfed group, length-for-age z-scores were significantly greater in all mixed-fed groups at 6 months of age *(P* ≤ 0.024 for all comparisons) and 12 months of age (*P* < 0.001 for all comparisons). Similarly, compared to the breastfed group, head circumference-for-age z-scores were significantly higher in the *B. lactis* and standard formula groups at 6 months of age (*P* ≤ 0.018) and in all mixed-fed groups at 12 months of age (*P* ≤ 0.006). As shown in Figure 2, the mean z-scores of each study group were below the WHO median for nearly the entire study duration, only approaching the median at the 12-month time point.

**Figure 2.**
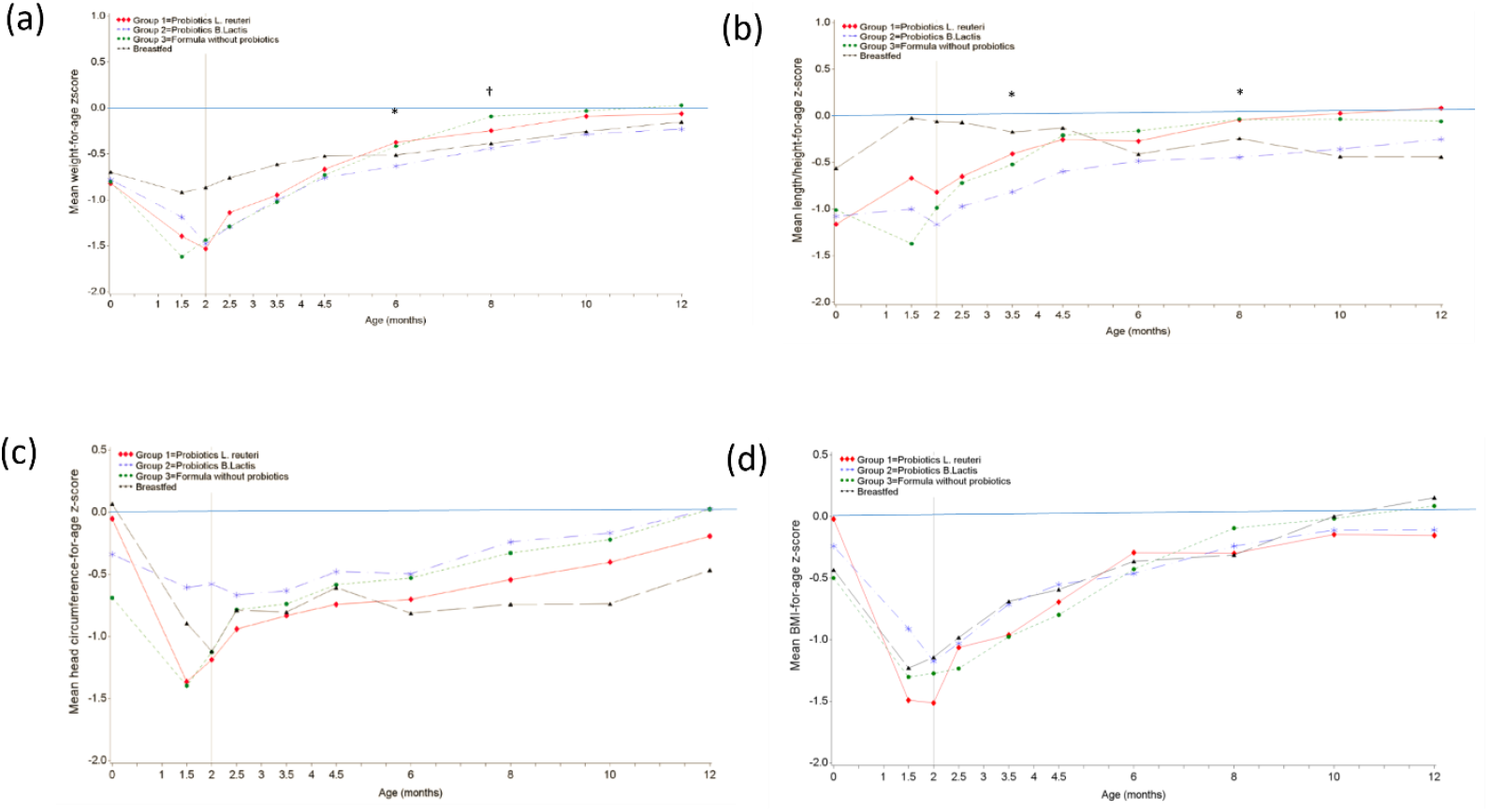
Mean weight gain-by-age Z-scores (a), length-for-age Z-scores (b), head circumference-for-age z-scores (c), and BMI-for-age Z-scores (d) over time in infants up to 12 months of age. Mean values are raw values and are not adjusted for baseline values or sex. Vertical line represents mean age at initiation of study formula in randomized groups. \**P* < 0.05 for Group 1 (*L. reuteri)* vs Group 2 (*B. lactis*); †*P* < 0.05 for Group 2 (B. lactis) vs Group 3 (standard formula).

### Formula intake and digestive tolerance

The overall number of breastfeeds per day were similar in the 3 formula groups (4.8 ± 2.6, 5.0 ± 2.4, and 5.0 ± 2.4 for the *L. reuteri, B. lactis*, and standard formula groups, respectively) and lower than the breastfed group (7.9 ± 0.31). Similarly, the overall reported formula intake per day was also similar in the 3 formula groups (505 ± 265, 487 ± 280, and 486 ± 241 mL/day for the *L. reuteri, B. lactis*, and standard formula groups, respectively). No formula intake was reported for the breastfed group until infants were aged 12 months, when average intake of 202 ± 243 mL/day was reported. Mean daily stool count was approximately 2 in all groups and approximately 80% to 90% of stools were either formed or soft (**Figure 3**). The number of days with reported frequent flatulence, frequent spitting up, or with at least 1 vomiting episode were similar in all feeding groups. Infants were reported as irritable and fussy approximately 20% of the time in all groups, with no significant differences between groups (**Table 2**).

**Table 2.**
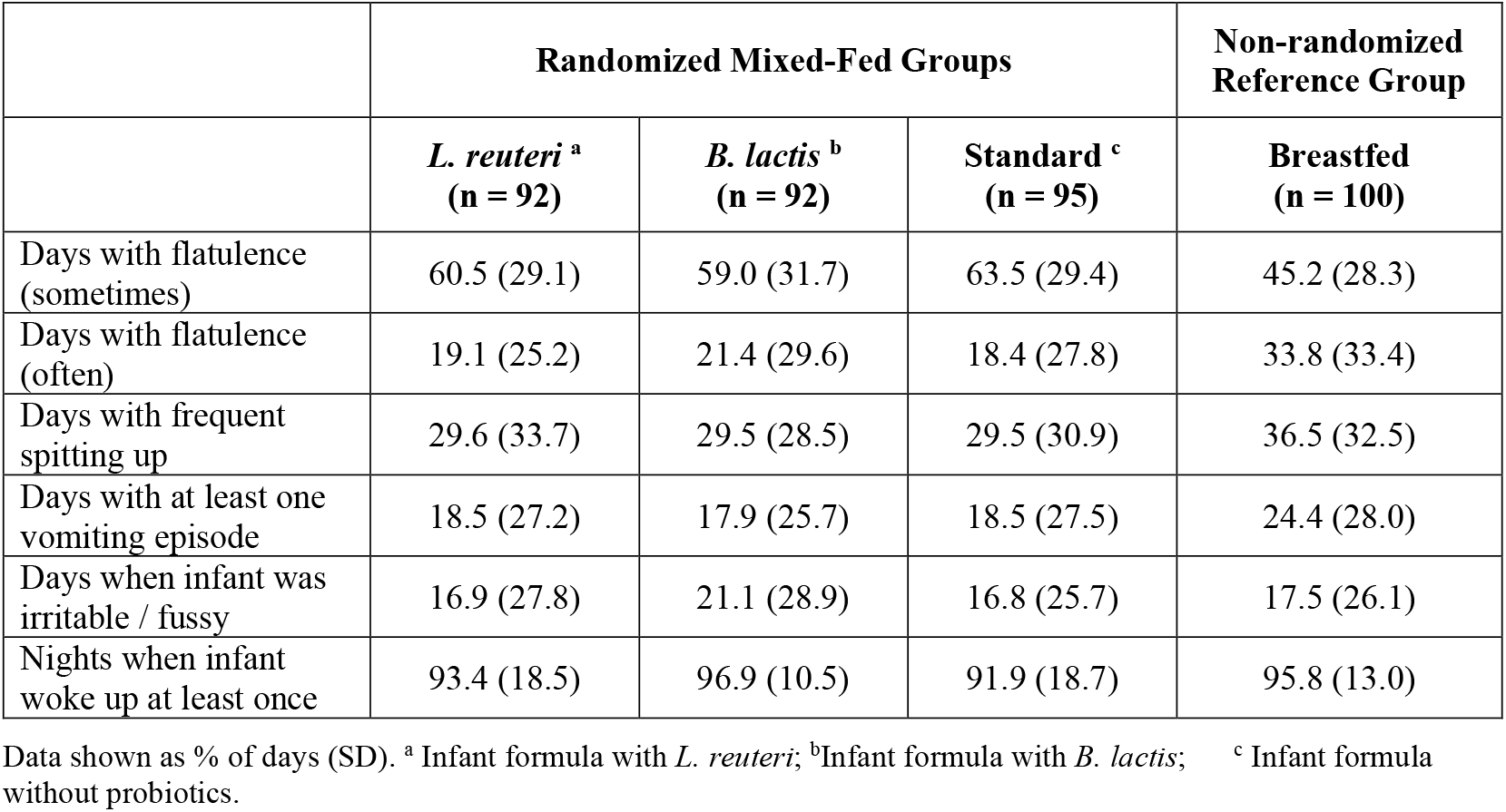
GI tolerance from randomization to age 6 months.

|  | Randomized Mixed-Fed Groups |  |  | Non-randomized Reference Group |
| --- | --- | --- | --- | --- |
|  | *L. reuteri*<sup>a</sup><br>(n = 92) | *B. lactis*<sup>b</sup><br>(n = 92) | Standard<sup>c</sup><br>(n = 95) | Breastfed<br>(n = 100) |
| Days with flatulence (sometimes) | 60.5 (29.1) | 59.0 (31.7) | 63.5 (29.4) | 45.2 (28.3) |
| Days with flatulence (often) | 19.1 (25.2) | 21.4 (29.6) | 18.4 (27.8) | 33.8 (33.4) |
| Days with frequent spitting up | 29.6 (33.7) | 29.5 (28.5) | 29.5 (30.9) | 36.5 (32.5) |
| Days with at least one vomiting episode | 18.5 (27.2) | 17.9 (25.7) | 18.5 (27.5) | 24.4 (28.0) |
| Days when infant was irritable / fussy | 16.9 (27.8) | 21.1 (28.9) | 16.8 (25.7) | 17.5 (26.1) |
| Nights when infant woke up at least once | 93.4 (18.5) | 96.9 (10.5) | 91.9 (18.7) | 95.8 (13.0) |

Data shown as % of days (SD). <sup>a</sup> Infant formula with *L. reuteri*; <sup>b</sup> Infant formula with *B. lactis*; <sup>c</sup> Infant formula without probiotics.

**Figure 3.**
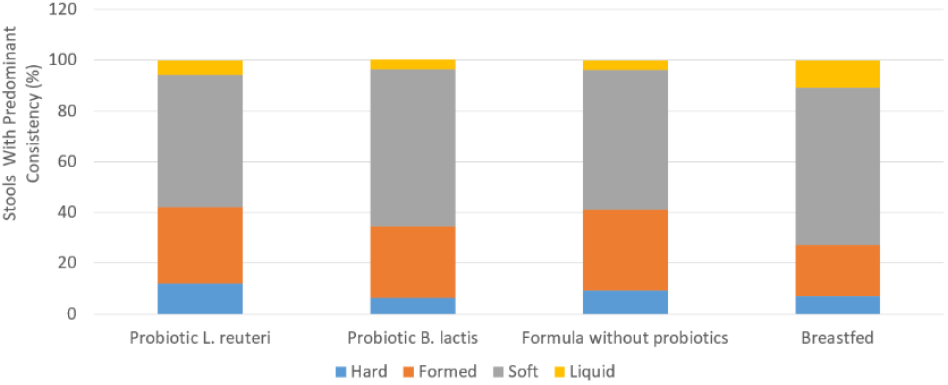
Total percentage of stools with their predominant stool consistency (hard, formed, soft, liquid) per group from randomization through month 6.

### Stool microbiota composition

The composition of stool microbiota was evaluated in a subset of infants at 4.5 months of age (**Table 3**). There were no statistically significant differences between either the *L. reuteri* or the *B. lactis* group compared to the standard formula group with regard to total bacteria, bifidobacteria, or Clostridium, Enterobacteriaceae, and Bacteroides counts. Lactobacillus cluster counts (particularly *L. reuteri* and *L. gasseri*) were significantly higher in the *L. reuteri* group compared to infants fed *B. lactis* (*P* = 0.005) or standard formula (*P* = 0.001) and compared to breastfed infants (*P* = 0.005). Breastfed infants had significantly higher total bacteria and bifidobacteria counts and significantly lower Clostridium counts compared to mixed-fed infants *(P ≤* 0.033 for all comparisons). Enterobacteriaceae counts with breastfed infants appeared similar to those in the *L. reuteri* and the standard formula groups and lower than those in the *B. lactis* group (*P* = 0.012).

**Table 3.** Infant stool microbiota composition in a subgroup of infants at 4.5 months of age.

|  | Randomized Mixed-Fed Groups |  |  | Non-randomized Reference Group |
| --- | --- | --- | --- | --- |
|  | *L. reuteri*<sup>a</sup><br>(n = 24) | *B. lactis*<sup>b</sup><br>(n = 25) | Standard<sup>c</sup><br>(n = 21) | Breastfed<br>(n = 30) |
| Total bacteria | 10.23 ± 0.45<sup>*</sup> | 10.27 ± 0.45<sup>*</sup> | 10.24 ± 0.52<sup>*</sup> | 10.60 ± 0.33<sup>†</sup> |
| Bifidobacteria | 9.30 ± 1.43<sup>*</sup> | 9.67 ± 0.92<sup>*</sup> | 9.21 ± 1.58<sup>*</sup> | 10.38 ± 0.90<sup>†</sup> |
| Clostridium / Eubacterium | 8.27 ± 1.50<sup>*</sup> | 7.57 ± 1.40<sup>*</sup> | 8.42 ± 1.60<sup>*</sup> | 6.70 ± 1.30<sup>†</sup> |
| Lactobacillus cluster | 7.01 ± 1.00<sup>*</sup> | 6.27 ± 0.71<sup>†</sup> | 6.18 ± 0.62<sup>†</sup> | 6.29 ± 0.65<sup>†</sup> |
| Enterobacteriaceae | 7.53 ± 1.29<sup>*</sup> | 8.24 ± 0.94<sup>†</sup> | 7.88 ± 1.22<sup>*†</sup> | 7.43 ± 1.20<sup>*</sup> |
| Bacteroides / Prevotella | 6.34 ± 0.96<sup>*</sup> | 6.08 ± 0.33<sup>*</sup> | 6.40 ± 0.88<sup>*</sup> | 6.05 ± 0.24<sup>*</sup> |

Mean ± SD bacterial count (log<sub>10</sub>[CFU/g]); values within a row with different superscript symbols are significantly different. <sup>a</sup> Infant formula with *L. reuteri*; <sup>b</sup> Infant formula with *B. lactis*; <sup>c</sup> Infant formula without probiotics.

### Urinary D-lactate

Urinary lactate levels, including D-lactate, L-lactate, percentage of D-lactate/L-lactate, and sum of D-lactate and L-lactate, were assessed in a subset of infants at approximately 2.5 months of age (2 months after the start of formula feeding in mixed-fed infants). No statistically significant differences were observed between any of the randomized formula groups or between the mixed-fed and the breastfed groups (**Table 4**).

**Table 4.** Urinary D-lactate parameters in a subgroup of infants at 2.5 months of age.

|  | Randomized Mixed-Fed Groups |  |  | Non-randomized Reference Group |
| --- | --- | --- | --- | --- |
|  | *L. reuteri*<sup>a</sup><br>(n = 10) | *B. lactis*<sup>b</sup><br>(n = 16) | Standard<sup>c</sup><br>(n = 14) | Breastfed<br>(n = 29) |
| D-lactic acid, mmol / mol creatinine | 7.9 ± 11.9 | 4.3 ± 7.7 | 6.7 ± 9.2 | 7.3 ± 11.3 |
| L-lactic acid, mmol / mol creatinine | 131 ± 210 | 269 ± 587 | 84 ± 110 | 152 ± 193 |
| Sum of D-lactate / L-lactate mmol / mol creatinine | 139.1 ± 215.3 | 273.3 ± 586.5 | 90.8 ± 117.3 | 159.1 ± 194.3 |
| D-lactate / L-lactate, % | 13.0 ± 22.8 | 7.8 ± 10.5 | 9.7 ± 11.1 | 9.9 ± 15.9 |

Mean ± SD; values within a row were not significantly different. <sup>a</sup>Infant formula with *L. reuteri*; <sup>b</sup>Infant formula with *B. lactis*; <sup>c</sup>Infant formula without probiotics.

### Adverse events

All infants who initiated formula (in the randomized groups) or entered into the study (breastfed groups) were included in the safety population. Treatment-emergent AEs occurred in 35 (38.0%) infants (110 events) in the *L. reuteri* group, 34 (37.0%) infants (100 events) in the *B. lactis* group, 41 (43.2%) infants (111 events) in the standard formula group, and 35 (35.0%) of infants (99 events) in the breastfed group. Serious treatment-emergent AEs were reported in 3 infants (3.3%; 3 events) in the *L. reuteri* group, 8 infants (8.7%; 9 events) in the *B. lactis* group, 3 infants (3.2%; 3 events) in the standard formula group, and 2 infants (2.0%; 2 events) in the breastfed group. Rates of the most commonly reported treatment-emergent AEs were generally similar among feeding groups (**Table 5**). In *post-hoc* analyses, the incidence of AEs between 6 and 12 months was compared between the *L. reuteri* group and controls. Children in the control group had a higher likelihood of AEs gastroenteritis (OR 4.29 [1.12-18.24], *P* = 0.0286) and infections and infestations (OR 1.99 [0.96, 4.28], *P* = 0.0728) compared to those in the *L. reuteri* group. There were also slight trends for greater usage of antidiarrheal/intestinal anti-inflammatory/anti-infective medications (OR 2.59 [0.77, 9.19], *P* = 0.1637) and drugs for functional gastrointestinal disorders (OR 3.58 [0.76, 24.36], *P* = 0.1695) in the control group vs. the *L. reuteri* group. Of the 17 serious AEs reported in the trial, only one (gastroenteritis/lactose intolerance, which occurred in the *B. lactis* group) was considered probably formula-related by the investigator.

**Table 5.** Adverse events occurring from randomization until age 12 months.

|  | **Randomized Mixed-Fed Groups** |  |  | **Non-randomized Reference Group** |
| --- | --- | --- | --- | --- |
|  | *L. reuteri*<sup>a</sup><br>(n = 92) | *B. lactis*<sup>b</sup><br>(n = 92) | Standard<sup>c</sup><br>(n = 95) | Breastfed<br>(n = 100) |
| Respiratory tract infection | 9 (9.8) | 11 (12.0) | 17 (17.9) | 15 (15.0) |
| Gastroenteritis | 11 (12.0) | 15 (16.3) | 15 (15.8) | 9 (9.0) |
| Cough | 12 (13.0) | 9 (9.8) | 10 (10.5) | 5 (5.0) |
| Upper respiratory tract infection | 7 (7.6) | 6 (6.5) | 12 (12.6) | 11 (11.0) |
| Lower respiratory tract infection | 10 (10.9) | 5 (5.4) | 4 (4.2) | 10 (10.0) |
| Pyrexia | 7 (7.6) | 8 (8.7) | 9 (9.5) | 4 (4.0) |
| Diarrhea | 5 (5.4) | 3 (3.3) | 3 (3.2) | 9 (9.0) |
| Rhinitis | 2 (2.2) | 4 (4.3) | 7 (7.4) | 3 (3.0) |
| Vomiting | 5 (5.4) | 2 (2.2) | 3 (3.2) | 2 (2.0) |

Data shown as number of infants (%); only adverse events occurring in $\geq 5\%$ of infants in any group are presented; values within a row were not significantly different. <sup>a</sup>Infant formula and then follow-up formula with *L. reuteri*;
<sup>b</sup>Infant formula with *B. lactis* and then follow-up formula with *L. rhamnosus* LPR + *B. longum* BL999; <sup>c</sup>Infant and follow-up formulas without probiotics.

## 4. Discussion

In this study, infant growth as assessed by mean weight gain per day in infants assigned to a infant formula supplemented with *L. reuteri* or *B. lactis* was non-inferior to the standard infant formula without probiotic supplementation.

Analysis of secondary growth outcomes based on the WHO Standards demonstrated no statistically significant differences in any of the *z*-score values for growth parameters (including *z*-scores for weight-for-age, length-for-age, and head circumference-for-age) between either of the probiotic formula groups and the standard formula group when infants were 6 or 12 months of age. However, compared to breastfed infants, *z*-scores at 12 months of age for weight-for-age, length-for-age and head circumference-for-age were significantly higher in all the mixed-fed groups. Interestingly, all study groups, including the breastfed reference group, grew faster during the study period 9 weeks to 6 months compared to the WHO standard. According to the WHO Standards, mean weight gain during this interval is, on average, 18.33 g/day. Mean weight gain was higher than this WHO standard in all 4 groups, with a difference of 4.15 g/day observed in the *L. reuteri* group (*P* < 0.001), 2.74 g/day with *B. lactis* (*P* < 0.001), 3.22 g/day with the standard formula (*P* < 0.001) and 1.38 g/day in breastfed infants (*P* = 0.013). In a study of infant growth in 5 developing countries, the mean height-for-age *z*-score in India was −0.6 at birth and −1.5 at age 12 months, indicating that growth was slower in infants in India compared to WHO reference standards (Lundeen *et al*., 2014). In the current study, mean length-for-age z-scores in infants aged 12 months ranged from −0.7 in the breastfed group to 0.2 in the *L. reuteri* group. These data indicate that growth was slightly faster in all 4 study groups than previously reported averages in Indian infants, yet the close approximation of 12-month *z*-scores with the WHO median suggests that growth rates were appropriate over this interval. Overall, anthropometry data in infants assigned to formulas supplemented with *L. reuteri* through 12 months of age or *B. lactis* through 6 months of age and L. rhamnosus LPR + B. longum BL999 from 7 to 12 months of age support normal, age-appropriate infant growth through the first year of life.

Infant formulas with either *L. reuteri* or *B. lactis* supplementation were well tolerated. According to physicians’ reports, the incidence of AEs in infants in either probiotic group was comparable to the incidence in the standard formula. Parent-reported stool characteristics and GI symptoms assessed up to 6 months of age were similar among all three formula groups. In addition, urinary lactate levels were comparable in all tstudy groups, including the breastfed reference group, at 2.5 months of age. An increase in D-lactate-producing bacteria in the colon can lead to D-lactate acidosis, which is associated with neurological impairment (Kowlgi and Chhabra, 2015); thus, urinary D-lactate can be used as a safety biomarker for trials of infant formula supplemented with probiotics (Sanders *et al*., 2010). Urinary D-lactate values for individual subjects remained entirely within ranges previously reported in healthy infants (Haschke-Becher *et al*., 2000; Haschke-Becher *et al*., 2008), and well below the values associated with D-lactate excess (e.g., ~60 mmol/mol creatinine (Bongaerts *et al*., 2000)). It is also worth noting that, in a subset of infants at 4.5 months of age, significantly higher stool lactobacilli count was observed in the *L. reuteri* formula group vs. the standard formula group and the breastfed group. This finding indicates that supplementation with *L. reuteri* may help promote a healthy gut microbiome.

The safety and tolerability of *L. reuteri* supplementation in our trial was consistent with findings from two earlier studies evaluating the safety of infant formulas containing *L. reuteri* (Lee Le *et al*., 2015; Papagaroufalis *et al*., 2014). In a previous study of 88 healthy full term newborns who were randomly assigned to a infant formula with *L. reuteri* or a control formula without probiotics (Papagaroufalis *et al*., 2014), all anthropometric measurements and safety parameters up to 6 months were comparable between the two formula groups. Further, *Bifidobacterium* and lactobacilli were detected more frequently in the *L. reuteri* group vs. control (Papagaroufalis *et al*., 2014). Although we did not observe a higher fecal *Bifidobacterium* abundance in the present study, this may be due to our inclusion of mixed-fed infants versus exclusively formula-fed infants studied by Papagaroufalis et al. (Papagaroufalis *et al*., 2014), which may have limited our ability to detect differences between groups.

Various studies involving a collective total of more than 1000 infants with *L. reuteri* doses as high as 1.2 × 10^9^ CFU/day have consistently demonstrated the safety and tolerability of *L. reuteri* supplementation (Abrahamsson *et al*., 2009; Connolly *et al*., 2005; Indrio *et al*., 2011; Papagaroufalis *et al*., 2014; Savino *et al*., 2010; Savino *et al*., 2018; Savino *et al*., 2007; Tyrsin *et al*., 2024, Weizman *et al*., 2005). The totality of evidence demonstrates the safety and suitability of supplementing infants with the probiotic *L. reuteri* in supporting normal infant growth, now with this study, we show safety and suitability at a concentration consistent with the estimated daily intake of *L. reuteri* from infant formula feeding.

*B. lactis* is one of the most extensively studied probiotic bacteria in infant nutrition. Its safety and tolerability are well established, both in healthy infants and toddlers and in at risk groups such as preterm infants and infants from HIV positive mothers (Steenhout *et al*., 2009; Tremblay *et al*., 2023)}. An independent safety evaluation by the European Society for Paediatric Gastroenterology Hepatology and Nutrition concluded that *B. lactis*, alone or in combination, supports normal growth in healthy term infants and is not associated with adverse outcomes (Braegger *et al*., 2011). The beneficial effects of *B. lactis* supplementation that have been reported in previous trials are numerous, and include an increase in the bifidobacteria in the gut microbiota, as well as reduced incidence of diarrhea, rotavirus shedding, upper respiratory tract infections, diaper rash, and hard stools, and improved immune response (Chen *et al*., 2024; Dekker *et al*., 2022; Fukushima *et al*., 1998; Haschke *et al*., 1998; Hertiš Petek *et al*., 2022; Holscher *et al*., 2012; Langhendries *et al*., 1995; Phuapradit *et al*., 1999; Saavedra *et al*., 1998; Saavedra *et al*., 1994). In the present study, microbiota profile and GI tolerance in the *B. lactis* and standard formula groups were generally similar, perhaps due to the inclusion of mixed-fed infants (since breastmilk is also a strong influence on these endpoints).

One strength of the study is the number and location of the study sites (5 centers, each in a geographically distinct region of India), which help to support the generalizability of these results to the Indian infant population. Another strength is the inclusion of mixed-fed infants, a population that is often excluded from infant formula trials even though many mothers introduce formula feeding as they prepare to wean or as maternity leave ends. One limitation of the study was the use of fluorescent *in situ* hybridization, which is a less sensitive methodology for assessing fecal microbiota composition and abundance compared to more modern techniques. Another limitation was the lack of assessment of gut microbiota at 12 months of age, which could have allowed evaluation of the effect of probiotic enrichment of the study follow-up formulas.

## 5. Conclusions

The results from this trial have demonstrated that infant formulas with *L. reuteri* or *B. lactis* compared with the same formula without probiotics promote adequate weight gain (g/day) in infants up to 6 months of age. The primary growth data, in conjunction with the growth outcomes based on the WHO Standards data (i.e., *z*-score values for growth parameters) during the entire 12-month study period, also demonstrate that follow-up formulas with *L. reuteri* or *L. rhamnosus* LPR + B. longum BL999, respectively, fed from 7 to 12 months of age support age-appropriate normal infant growth. These findings confirm the results of similar evaluations of infant formulas supplemented with probiotics in different geographic locations and populations (Gibson *et al*., 2009; Lee Le *et al*., 2015; Papagaroufalis *et al*., 2014; Soh *et al*., 2009; Weizman *et al*., 2005). Further, a significantly higher lactobacilli count in the *L. reuteri* formula group vs. all other study groups, along with *Enterobacteriaceae* and *Bacteroides* / *Prevotella* counts that were similar to those of breastfed infants, indicate that supplementation with *L. reuteri* may help promote a healthy gut microbiome.

Rates of reported AEs with either the *L. reuteri*-supplemented formula or the *B. lactis* supplemented formula were comparable to those seen with both the standard infant formula and the breastfed reference group during the entire 12-month study period. The incidences of parent-reported GI symptoms such as spitting-up, vomiting, and irritability/fussiness were also similar among the 3 mixed-fed groups and comparable to those reported in the breastfed group. Consequently, this study confirms that infant formulas supplemented with probiotics *L. reuteri* or *B. lactis* are safe, well tolerated, and support normal infant growth in an Indian infant population.

## Data Availability

All gut microbiota data produced in the present study are available upon reasonable request to the authors.

## Acknowledgements

The authors thank the research staff at each institution and the families who volunteered to participate. We also thank Emilie Darcillon for assistance with trial management and Dominik Grathwohl, Mickaël Hartweg, Bridget-Anne Kirwan and Emille Perrin for assistance with statistical analysis. We also thank Sophie Pecquet for assistance with manuscript preparation.

The study was funded by Société des Produits Nestlé S.A. (previously Nestec Ltd). Co-authors employed by the funding body were involved in designing the study, data analysis and interpretation, and writing the manuscript.

## Author Contributions

MM, RA, UV, and SG conceived and designed the study. MM, RA, and UV recruited study participants and supervised data collection. All authors jointly contributed to analysis and interpretation of data. AP and NPH developed the first draft of the manuscript, with editorial support provided by Cooper Johnson Communications, LLC, funded by Nestlé. All authors reviewed manuscript drafts and approved the final version.

